# Assessment of the Modified Rankin Scale in Electronic Health Records with a Fine-tuned Large Language Model

**DOI:** 10.1101/2025.04.30.25326777

**Authors:** Luis Silva, Marcus Milani, Sohum Bindra, Salman Ikramuddin, Megan Tessmer, Kaylee Frederickson, Abhigyan Datta, Halil Ergen, Alex Stangebye, Dawson Cooper, Kompal Kumar, Jeremy Yeung, Kamakshi Lakshminarayan, Christopher D Streib

## Abstract

**Introduction:** The modified Rankin scale (mRS) is an important metric in stroke research, often used as a primary outcome in clinical trials and observational studies. The mRS can be assessed retrospectively from electronic health records (EHR), though this process is labor-intensive and prone to inter-rater variability. Large language models (LLMs) have demonstrated potential in automating clinical text classification. We hypothesize that a fine-tuned LLM can analyze EHR text and classify mRS scores for clinical and research applications.

**Methods:** We performed a retrospective cohort study of patients admitted to a specialist stroke neurology service at a large academic hospital system between August 2020 and June 2023. Each patient’s medical record was reviewed at two time points: (1) hospital discharge and (2) approximately 90 days post-discharge. Two independent researchers assigned an mRS score at each time point. Two separate models were trained on EHR passages with corresponding mRS scores as labeled outcomes: (1) a multiclass model to classify all seven mRS scores and (2) a binary model to classify functional independence (mRS 0–2) versus non-independence (mRS 3–6). Four-fold cross-validation was conducted, using accuracy and Cohen’s kappa as model performance metrics.

**Results:** A total of 2,290 EHR passages with corresponding mRS scores were included in model training. The multiclass model—considering all seven scores of the mRS—attained an accuracy of 77% and a weighted Cohen’s Kappa of 0.92. Class-specific accuracy was highest for mRS 4 (90%) and lowest for mRS 2 (28%). The binary model—considering only functional independence vs non-independence —attained an accuracy of 92% and Cohen’s Kappa of 0.84.

**Conclusion:** Our findings demonstrate that LLMs can be successfully trained to determine mRS scores through EHR text analysis. With further advancements, fully automated LLMs could scale across large clinical datasets, enabling data-driven public health strategies and optimized resource allocation.

## Introduction

The modified Rankin scale (mRS) is an important metric in stroke research, often used as a primary outcome in clinical trials and observational studies^1,2^. The mRS is scored from 0 (no symptoms) to 6 (death), with higher scores indicating greater disability. It is determined based on a patient’s stroke deficits and ability to perform daily activities^3^. It has also been used in pivotal stroke trials as a binary outcome comparing functional independence (scores 0-2) versus non-independence (scores 3-6)^4^. In most instances, trained clinicians or researchers collect the mRS in real-time. Alternatively, it can be assessed retrospectively from electronic health records (EHR), though this process is labor-intensive and prone to inter-rater variability^5–8^. These limitations constrain stroke research by making it dependent on the availability of trained research staff and prevents use of existing clinical databases for research.

Large language models (LLMs), such as GPT-4, are deep learning-based models that perform well in text classification and generation^9,10^. Use in medical research is expanding, with notable examples in neuroscience. GPT-4 has demonstrated 84% accuracy in localizing neurologic lesions and has also achieved a passing grade on the American Board of Psychiatry and Neurology examination^11,12^. However, GPT-4 performed poorly when assessing the Glasgow Coma Scale, the Intracranial Hemorrhage score, and the Hunt & Hess classifications^13^. These studies did not involve fine-tuning, a process in which the base model is trained on task-specific data to enhance performance. Classification of mRS scores from EHR text has been previously developed by Fernandez and colleagues^14^ who, using a non-LLM model, achieved 59% accuracy, limiting its practical application in both clinical and research settings.

We hypothesize that a fine-tuned LLM can analyze EHR text from inpatient and outpatient settings and classify mRS scores for clinical and research applications. We aimed to develop a tool capable of streamlining observational stroke research and reducing reliance on trained research staff.

## Methods

We performed a retrospective cohort study of patients evaluated by a stroke neurology service at a large academic hospital system between August 2020 and June 2023, following approval from the University of Minnesota Institutional Review Board. Each patient’s medical record was reviewed at two time points: (1) hospital discharge and (2) approximately 90 days post-discharge, with follow-up notes selected within a window of 30 to 120 days.

To minimize variability in follow-up timing, researchers were instructed to evaluate clinical notes recorded as close as possible to 90 days post-discharge. If no appropriate notes were available, they gradually expanded the search window in both directions, extending to a final range of 30 to 120 days post-discharge. At each time point, two independent researchers— trained and certified in mRS assessment—assigned an mRS score. mRS scoring followed the Rankin Focused Assessment, a structured checklist that standardizes patient evaluation through a question-answer format. Researchers assigned scores by answering predefined questions about functional status and stroke-related deficits^15^. Discrepancies in mRS scoring were resolved through discussion. If no consensus was reached, a third reviewer adjudicated the case.

Additionally, researchers identified and collected one corresponding EHR passage from the clinical note deemed critical for determining the mRS score. In order to prevent data leakage, direct mentions of mRS scores could not be included. This process generated the study’s observational unit: a paired EHR passage and mRS score. Because multiple, distinct EHR passages can support a single mRS score, each patient could contribute up to four EHR passage– mRS score pairs to the dataset: two from the discharge summary and two from the follow-up visit (one from each researcher at each timepoint). If the two researchers initially disagreed on the mRS score before reaching consensus, only the correctly adjudicated mRS score-EHR passage was included in the data set. If a consensus could not be reached on the mRS score, both observational units were excluded from the analysis. Figure 1a illustrates EHR text collection, mRS scoring, and data inclusion decisions for model training.

**Figure 1.**
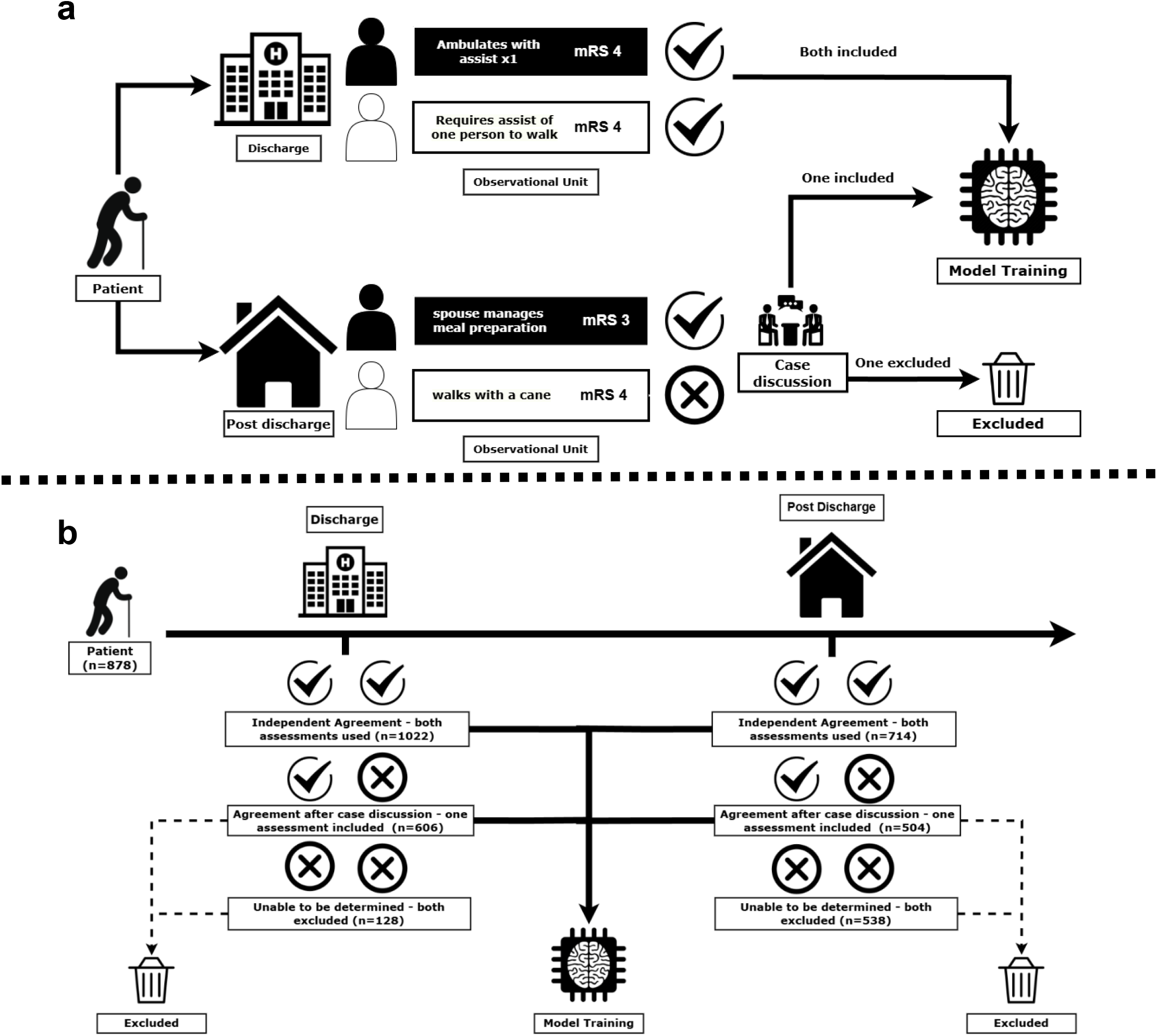
(a) Illustrative example of the data inclusion and exclusion process for each potential observational unit of a hypothetical patient. (b) Actual data inclusion and exclusion for LLM training within our study cohort

To develop our model, we used an existing LLM as the basis—Gatortron. Created at the University of Florida, Gatortron was trained using real-world EHR notes from the UFHealth system^16^. This allows it to capture the nuances of EHR text, making it suitable for our research. We fine-tuned Gatortron on our dataset to classify patients’ mRS scores based on the paired EHR passage. Fine-tuning is the process of further training a pre-trained language model on a specific task—in this case, classifying mRS scores from EHR passages—to enable the model to adapt its general language understanding to the nuances of functional outcome assessment in stroke^17^. Two separate models were trained: (1) a multiclass model to classify all seven mRS scores and (2) a binary model to classify functional independence (mRS 0–2) versus non-independence (mRS 3–6). Four-fold cross-validation was conducted, using accuracy, unweighted, and weighted Cohen’s kappa as performance metrics. EHR passage collected by researchers was used in its original form, without additional preprocessing or normalization.

During training, mRS classes were assigned training weights based on their overall frequency in the dataset—lower-frequency classes had proportionally higher weights. This approach balanced the dataset, reducing the likelihood of the model overfitting to more common cases.

Additionally, each fold was stratified to prevent model overfitting. Appendix 1 contains detailed model information.

The data supporting this study’s findings are available from the corresponding author upon reasonable request. The University of Minnesota—Twin Cities Institutional Review Board approved this research

## Results

878 patients were evaluated, contributing 2,290 observational units (EHR passage-mRS score pairs) to LLM training. Observational units were distributed across two stages: discharge (n=1,325) and 90 days post-discharge (n=966). This represents 75% (1,325/1,765) of the potential observational units at discharge and 55% (966/1,765) of the potential observational units from the post-discharge period. Figure 1b details the process of EHR passage-mRS score pairs inclusion and exclusion at each time point. The demographics were similar between groups, with a median age of 71 years (IQR 60–81) vs. 70 years (IQR 59–80), ischemic stroke proportions of 84% vs. 87%, and hemorrhagic stroke proportions of 10% vs. 8.2% (Table 1). The median time from hospitalization to discharge was 1 day (IQR 1–3), and from hospitalization to follow-up was 79 days (IQR 56–103).

**Table 1.**
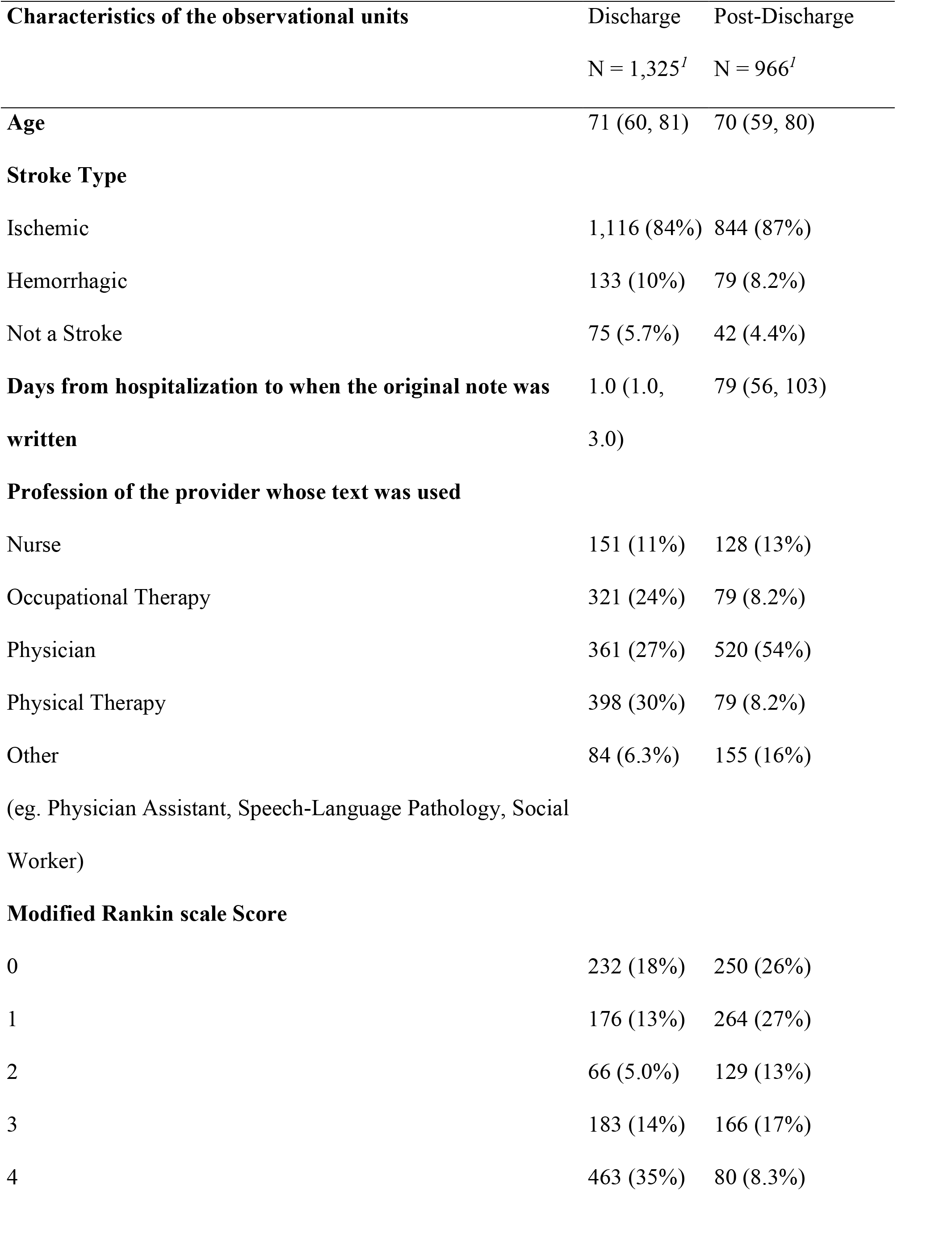
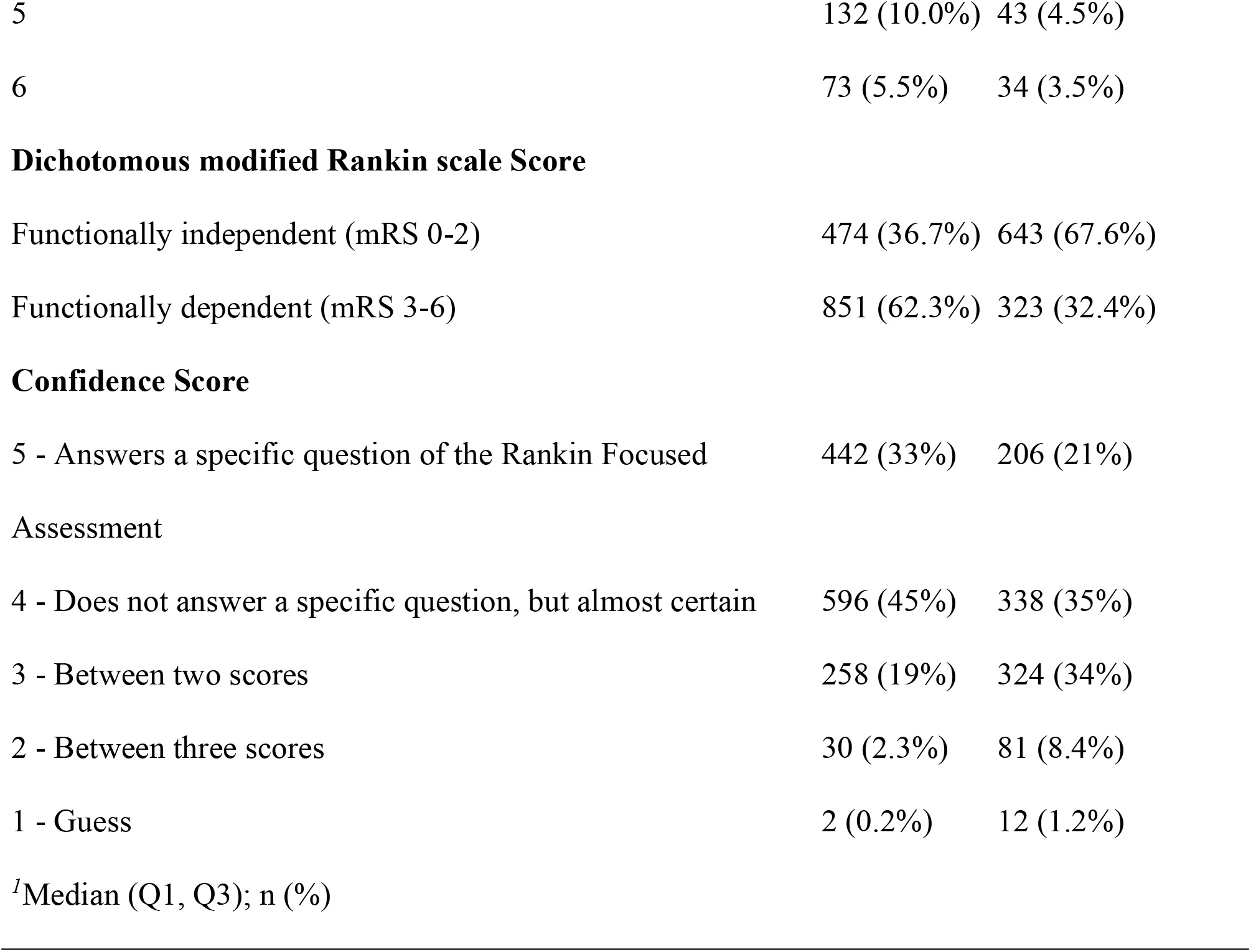
Model training and evaluation data characteristics.

| **Characteristics of the observational units** | Discharge | Post-Discharge |
| --- | --- | --- |
|  | N = 1,325<sup>I</sup> | N = 966<sup>I</sup> |
| **Age** | 71 (60, 81) | 70 (59, 80) |
| **Stroke Type** |  |  |
| Ischemic | 1,116 (84%) | 844 (87%) |
| Hemorrhagic | 133 (10%) | 79 (8.2%) |
| Not a Stroke | 75 (5.7%) | 42 (4.4%) |
| **Days from hospitalization to when the original note was written** | 1.0 (1.0, 3.0) | 79 (56, 103) |
| **Profession of the provider whose text was used** |  |  |
| Nurse | 151 (11%) | 128 (13%) |
| Occupational Therapy | 321 (24%) | 79 (8.2%) |
| Physician | 361 (27%) | 520 (54%) |
| Physical Therapy | 398 (30%) | 79 (8.2%) |
| Other<br>(eg. Physician Assistant, Speech-Language Pathology, Social Worker) | 84 (6.3%) | 155 (16%) |
| **Modified Rankin scale Score** |  |  |
| 0 | 232 (18%) | 250 (26%) |
| 1 | 176 (13%) | 264 (27%) |
| 2 | 66 (5.0%) | 129 (13%) |
| 3 | 183 (14%) | 166 (17%) |
| 4 | 463 (35%) | 80 (8.3%) |
| 5 | 132 (10.0%) | 43 (4.5%) |
| 6 | 73 (5.5%) | 34 (3.5%) |
| **Dichotomous modified Rankin scale Score** |  |  |
| Functionally independent (mRS 0-2) | 474 (36.7%) | 643 (67.6%) |
| Functionally dependent (mRS 3-6) | 851 (62.3%) | 323 (32.4%) |
| **Confidence Score** |  |  |
| 5 - Answers a specific question of the Rankin Focused Assessment | 442 (33%) | 206 (21%) |
| 4 - Does not answer a specific question, but almost certain | 596 (45%) | 338 (35%) |
| 3 - Between two scores | 258 (19%) | 324 (34%) |
| 2 - Between three scores | 30 (2.3%) | 81 (8.4%) |
| 1 - Guess | 2 (0.2%) | 12 (1.2%) |

<sup>1</sup>Median (Q1, Q3); n (%)

The two-class model, which combined the mRS scores into two categories (mRS 0-2 vs. mRS 3-6), had an accuracy of 92% and a Cohen’s Kappa of 0.85, and the model’s confusion matrix is shown in Figure 2. The multi-class model, which included all seven categories of the mRS, achieved an accuracy of 77%, a Cohen’s Kappa of 0.71, and a weighted Kappa of 0.92. Figure 3 represents the confusion matrix for this model and shows that the highest classification accuracy occurred for scores of 0 (90%) and 6 (99%). Misclassification was more common among intermediate scores, particularly score 2, where class accuracy was limited to 25%. Greater variability was observed in the most common words for intermediate mRS scores versus extreme scores, which complicated their classification. This finding is illustrated by Figure 4, a heatmap of the most common words in each mRS strata.

**Figure 2.**
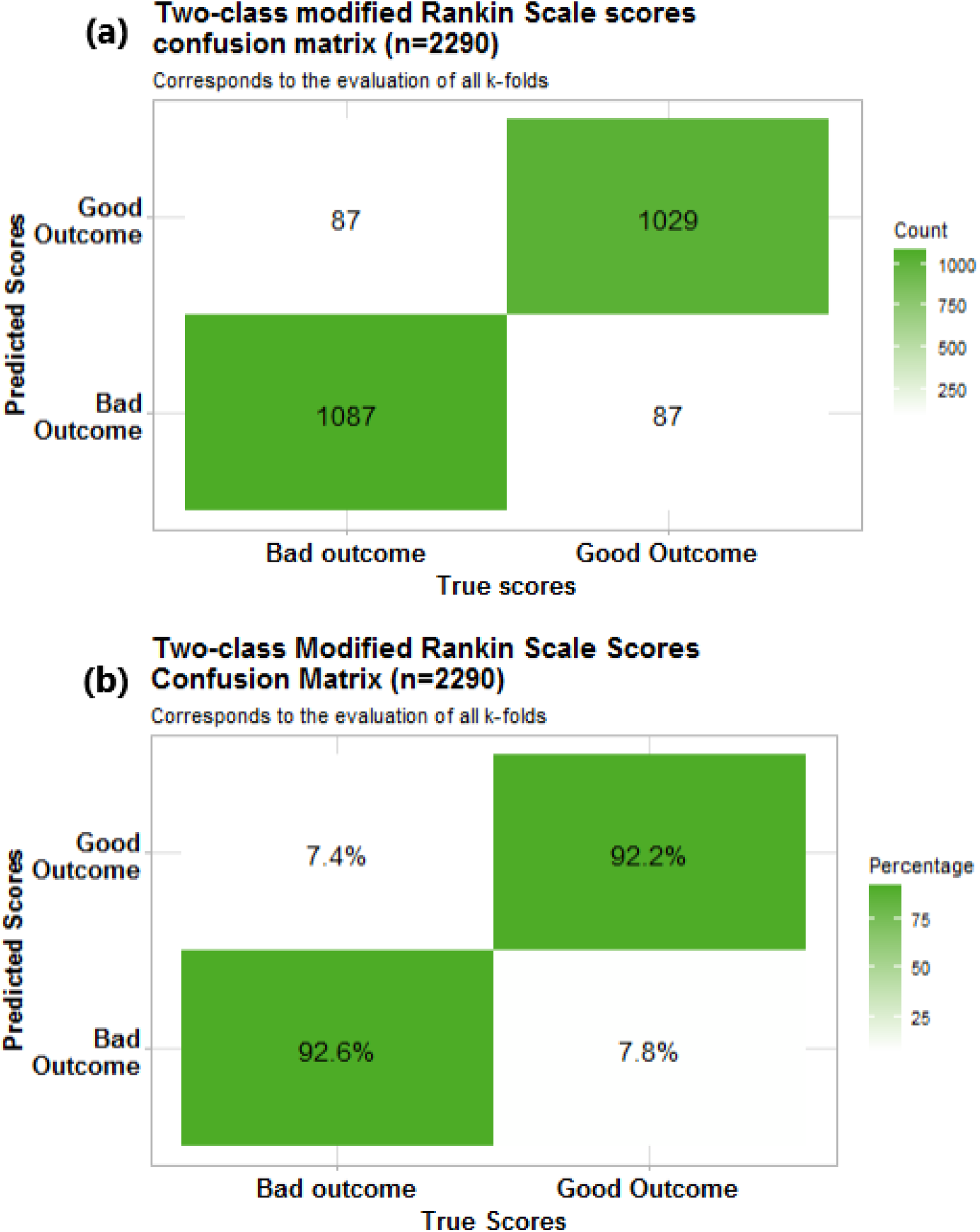
Confusion Matrix of the binomial classification model as (a) absolute counts and (b) class accuracy

**Figure 3.**
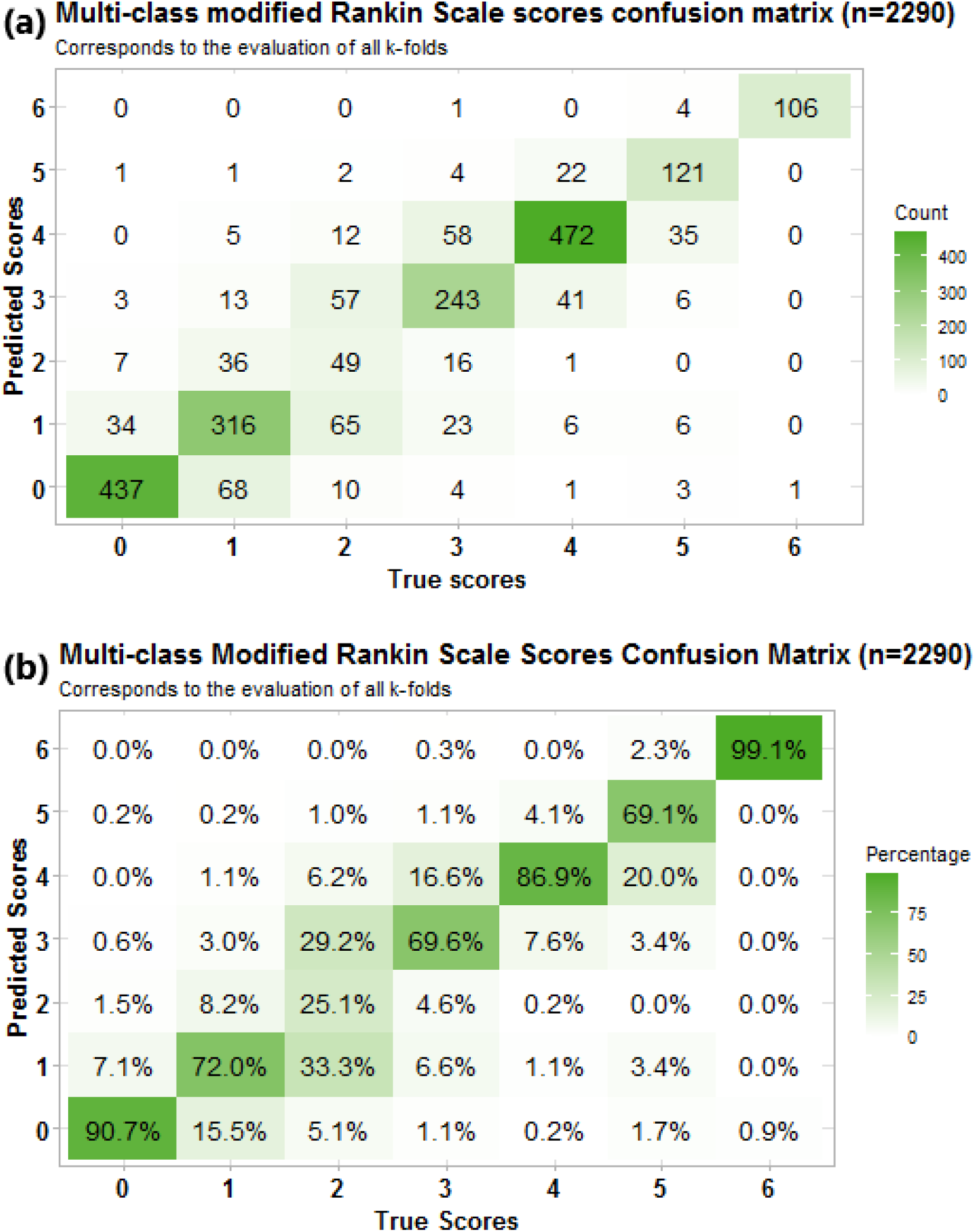
Confusion Matrix of the multiclass classification model as (a) absolute counts and (b) class accuracy

**Figure 4.**
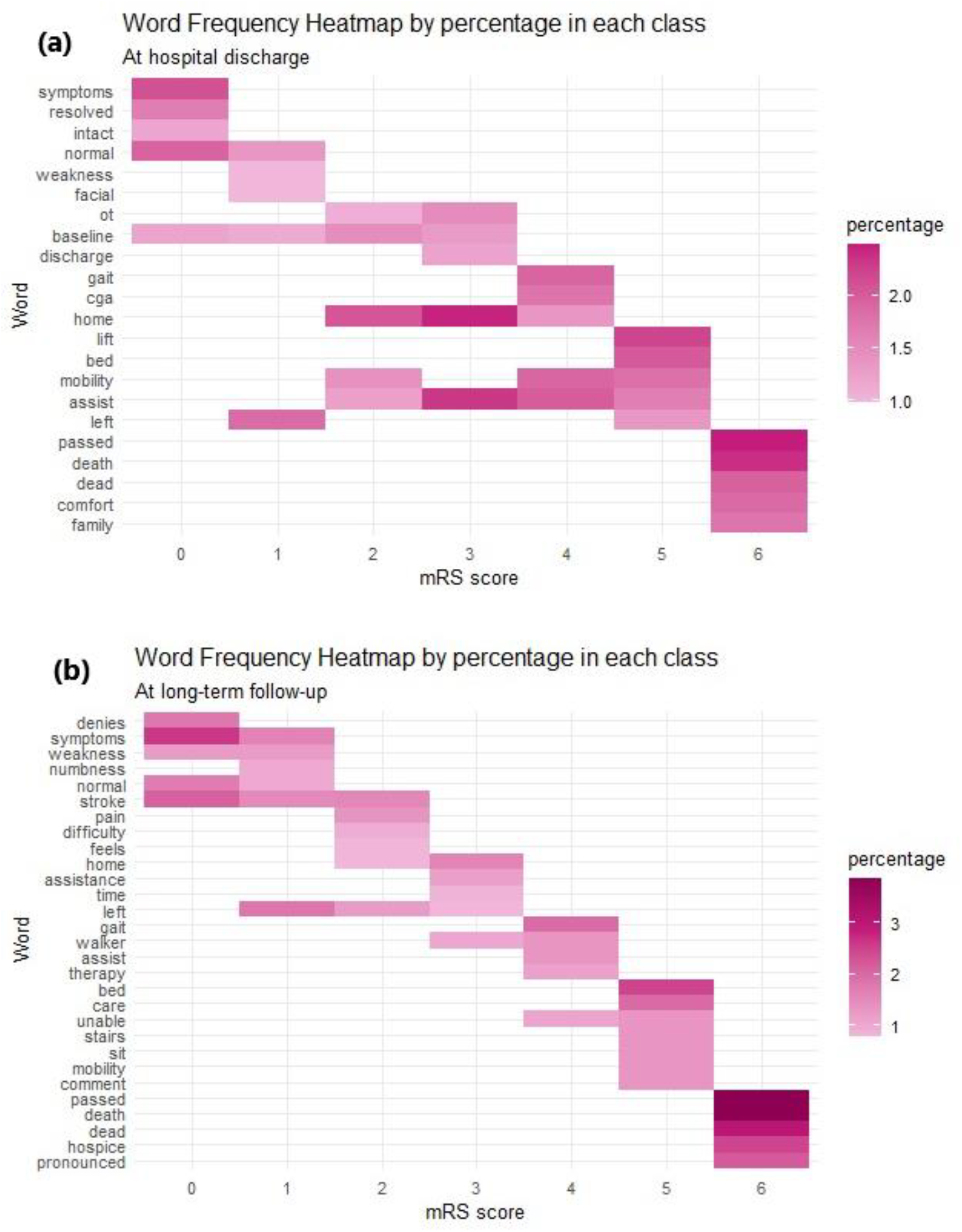
Heatmap of the most common words extracted in EHR passage for each class at (a) discharge and (b) long-term follow-up.

## Discussion

The mRS has become a standard outcome measure in stroke clinical trials due to its ability to capture significant levels of disability while remaining intuitive and less time-intensive to ascertain compared to more detailed metrics^15,18^. Automated tools capable of deriving the mRS from EHRs could streamline clinical research and potentially enable large-scale analyses using existing databases that lack mRS outcome data.

Our multiclass model demonstrated an accuracy of 77%, marking a significant improvement over previously published machine learning models for mRS determination from the EHR, which reported an accuracy of 59%^14^. Furthermore, our model also had a Cohen’s Kappa of 0.71 and a weighted Kappa of 0.92. The weighted Kappa, which penalizes larger classification errors more heavily than smaller ones, further underscores the model’s performance by indicating that most errors are near-misses rather than substantial misclassifications. In a systematic review of stroke trials, human raters classified mRS with an average agreement accuracy of 73%, unweighted Kappa of 0.62, weighted Kappa of 0.87^6^. This suggests our model performs comparably to human raters and could assist in real-world applications.

Our two-class model achieved an accuracy of 92% in differentiating patients who achieved functional independence (mRS score 0-2) from those with functional dependence (mRS score 3-6). This demonstrates that reducing the number of classes can significantly improve model performance, a key consideration depending on application.

We observed poorer model performance in the multiclass model, especially when differentiating the mRS score 2, and 3 (Figure 3). The model’s lower class-specific accuracies for intermediate mRS scores illustrate the inherent challenges of assessing mRS. In a study of 7,374 patients, mRS scores of 0–2 showed narrow variability in Longshi scale and Barthel Index scores, whereas mRS scores of 3–4 exhibited much broader variability^19^. This disparity demonstrates that while the mRS score effectively identifies stroke patients with mild disability, it is less capable of differentiating between moderate and severe disability. This known limitation of the mRS contributed to our choice of study design, which included only instances in which score consensus was achieved and excluded cases where EHR notes were ambiguous regarding a patient’s functional status. While this design helped maintain a reliable dataset, we acknowledge that there is likely selection bias due to the exclusion of cases that may be common in clinical practice. Future iterations of LLMs should assess performance across sociodemographic groups, representing an important area for future investigation.

Rather than replacing human raters, our model could assist in mRS assessment by confirming initial evaluations. We propose that the model could act as a secondary reviewer flagging uncertain cases for additional human review. By providing the model with passages of EHR text, independent confirmation of mRS scores can occur, using an additional reviewer only for complex cases. This approach could reduce the number of personnel needed for chart review studies while also improving the reproducibility of data collection.

With minor prompt engineering, our model could be trained to output class probabilities rather than discrete scores, allowing users to change the threshold of class determination based on the model application. With further advancements, fully automating mRS scoring from unstructured clinical text could integrate real-time outcome metrics into Learning Health Systems. This enables dynamic feedback to guide healthcare delivery and system-level decision-making. Our findings support the continued validation, investigation, and integration of LLMs into medical research and clinical care

## Conclusions

Our study found that a fine-tuned LLM using EHR passages could accurately classify 77% of mRS scores in the multiclass model (scoring mRS 0-6) and 92% of mRS scores in the two-class model (stratifying mRS 0-2 versus mRS 3-6). Utilizing fine-tuned LLMs to determine mRS scores could streamline data analysis for research and clinical applications.

## Data Availability

The data that support the findings of this study are available from the corresponding author upon reasonable request.

## Non-standard Abbreviations and Acronyms

(EHR): Electronic Health Records
(LLM): Large Language Model
(mRS): Modified Rankin Scale

## Acknowledgments

The authors would like to thank the Department of Neurology and the School of Public Health at the University of Minnesota for their invaluable efforts in making this research possible.

Additionally, we would like to acknowledge the use of automated assistive writing technologies and tools in manuscript and coding development. They have been used as supportive adjuncts to refine the main structure of both aspects of this research.

## References

1. Li Q, Abdalkader M, Siegler JE, Yaghi S, Sarraj A, Campbell BCV, Yoo AJ, Zaidat OO, Kaesmacher J, Pujara D, et al. Mechanical Thrombectomy for Large Ischemic Stroke: A Systematic Review and Meta-analysis. Neurology. 2023;101:e922–e932.

2. Hendrix P, Collins MK, Griessenauer CJ, Goren O, Melamed I, Weiner GM, Dalal SS, Kole MJ, Noto A, Schirmer CM. Tenecteplase versus alteplase before mechanical thrombectomy: experience from a US healthcare system undergoing a system-wide transition of primary thrombolytic. J. NeuroInterventional Surg. 2022;jnis-2022-019662.

3. Broderick JP, Adeoye O, Elm J. The Evolution of the Modified Rankin Scale and Its Use in Future Stroke Trials. Stroke. 2017;48:2007–2012.

4. Berkhemer OA, Fransen PSS, Beumer D, Berg LA van den, Lingsma HF, Yoo AJ, Schonewille WJ, Vos JA, Nederkoorn PJ, Wermer MJH, et al. A Randomized Trial of Intraarterial Treatment for Acute Ischemic Stroke. N. Engl. J. Med. 2015;372:11–20.

5. Sucharew H, Kleindorfer D, Khoury JC, Alwell K, Haverbusch M, Stanton R, Demel S, De Los Rios La Rosa F, Ferioli S, Jasne A, et al. Deriving Place of Residence, Modified Rankin Scale, and EuroQol-5D Scores from the Medical Record for Stroke Survivors. Cerebrovasc. Dis. Basel Switz. 2021;50:567–573.

6. Quinn TJ, Dawson J, Walters MR, Lees KR. Reliability of the Modified Rankin Scale. Stroke. 2009;40:3393–3395.

7. Banks JL, Marotta CA. Outcomes Validity and Reliability of the Modified Rankin Scale: Implications for Stroke Clinical Trials. Stroke. 2007;38:1091–1096.

8. Zhao H, Collier JM, Quah DM, Purvis T, Bernhardt J. The Modified Rankin Scale in Acute Stroke Has Good Inter-Rater-Reliability but Questionable Validity. Cerebrovasc. Dis. 2009;29:188–193.

9. Vaswani A, Shazeer N, Parmar N, Uszkoreit J, Jones L, Gomez AN, Kaiser L, Polosukhin Attention Is All You Need [Internet]. 2023 [cited 2024 Sep 21];Available from: http://arxiv.org/abs/1706.03762

10. OpenAI, Achiam J, Adler S, Agarwal S, Ahmad L, Akkaya I, Aleman FL, Almeida D, Altenschmidt J, Altman S, et al. GPT-4 Technical Report [Internet]. 2024 [cited 2024 Sep 21];Available from: http://arxiv.org/abs/2303.08774

11. Lee J-H, Choi E, McDougal R, Lytton WW. GPT-4 Performance for Neurologic Localization. Neurol. Clin. Pract. 2024;14:e200293.

12. Schubert MC, Wick W, Venkataramani V. Performance of Large Language Models on a Neurology Board–Style Examination. JAMA Netw. Open. 2023;6:e2346721.

13. Chen TC, Kaminski E, Koduri L, Singer A, Singer J, Couldwell M, Delashaw J, Dumont A, Wang A. Chat GPT as a Neuro-Score Calculator: Analysis of a Large Language Model’s Performance on Various Neurological Exam Grading Scales. World Neurosurg. 2023;179:e342–e347.

14. Fernandes MB, Valizadeh N, Alabsi HS, Quadri SA, Tesh RA, Bucklin AA, Sun H, Jain A, Brenner LN, Ye E, et al. Classification of neurologic outcomes from medical notes using natural language processing. Expert Syst. Appl. 2023;214:119171.

15. Saver JL, Filip B, Hamilton S, Yanes A, Craig S, Cho M, Conwit R, Starkman S. IMPROVING THE RELIABILITY OF STROKE DISABILITY GRADING IN CLINICAL TRIALS AND CLINICAL PRACTICE: THE RANKIN FOCUSED ASSESSMENT (RFA). Stroke J. Cereb. Circ. 2010;41:992–995.

16. Yang X, Chen A, PourNejatian N, Shin HC, Smith KE, Parisien C, Compas C, Martin C, Flores MG, Zhang Y, et al. GatorTron: A Large Clinical Language Model to Unlock Patient Information from Unstructured Electronic Health Records [Internet]. 2022 [cited 2025 Feb 18];Available from: http://arxiv.org/abs/2203.03540

17. Wang A, Liu C, Yang J, Weng C. Fine-tuning large language models for rare disease concept normalization. J. Am. Med. Inform. Assoc. 2024;31:2076–2083.

18. Quinn TJ, Dawson J, Walters MR, Lees KR. Functional outcome measures in contemporary stroke trials. Int. J. Stroke Off. J. Int. Stroke Soc. 2009;4:200–205.

19. Kwon S, Hartzema AG, Duncan PW, Min-Lai S. Disability measures in stroke: relationship among the Barthel Index, the Functional Independence Measure, and the Modified Rankin Scale. Stroke. 2004;35:918–923.

